# Relative wealth and inequality associate with health in a small-scale subsistence society

**DOI:** 10.1101/2020.06.11.20121889

**Authors:** Adrian V. Jaeggi, Aaron D. Blackwell, Christopher von Rueden, Benjamin C. Trumble, Jonathan Stieglitz, Angela R. Garcia, Thomas S. Kraft, Hillard Kaplan, Michael Gurven

## Abstract

In high-income countries, relative wealth and inequality may affect health by causing psychosocial stress. We test this hypothesis in a small-scale subsistence society, the Tsimane. We associated relative household wealth (n=1003) and community-level wealth inequality (n=35, Gini=0.15 – 0.43) with a range of psychosocial and health outcomes (depressive symptoms [n=663], social conflicts [n=393], nonsocial problems [n=390], social support [n=392], cortisol [n=828], BMI [n=9378], blood pressure [n=1614]), self-rated health [n=809], morbidities [n=3140]) controlling for absolute wealth, age, sex, community size, distance to town and relevant random effects. Relative wealth and inequality were associated with self-rated health and morbidity, especially respiratory disease, the leading cause of mortality in the Tsimane. Inequality was also associated with higher blood pressure. However, psychosocial stress did not mediate these associations, suggesting other mechanisms. These findings are consistent with socio-economic hierarchies affecting some health outcomes in any society, while others might be exacerbated in high-income countries.

## Introduction

It is relatively uncontroversial that people with greater access to resources – usually operationalized as income, wealth, or broader indicators of socio-economic position, rank or status^1^ – should be in better health, as resources can be converted into better nutritional status, access to health care, or insulation against health risks. Such benefits of *absolute* rank are also commonly found in non-human primates (Cowlishaw and Dunbar, 1991; Pusey et al., 1997; Snyder-Mackler et al., 2020; van Noordwijk and van Schaik, 1999). However, there is increasing evidence that *relative* access to resources, i.e. one’s relative position in a socio-economic hierarchy may also affect health. Across developed societies, there is causal evidence for a health gradient along socio-economic hierarchies, independent of absolute wealth or use of health care services (Ecob and Davey Smith, 1999; Marmot et al., 1991; Oakes et al., 1973; Sorlie et al., 1995; Wolfson et al., 1993). In other words, these studies find that *relative* rank – how one compares to others – is a critical variable in determining health outcomes (Anderson et al., 2012; Luttmer, 2005; Snyder-Mackler et al., 2020; Wood et al., 2012).

Furthermore, studies have found that the steepness of socio-economic hierarchies (i.e. income or wealth *inequality*) is associated with both physical and mental health outcomes – including self-rated health, all-cause mortality, heart disease, respiratory disease, obesity, or homicide – independent of the effects of absolute wealth (Nowatzki, 2012; Pickett and Wilkinson, 2015; Wilkinson and Pickett, 2006). While these findings are hotly debated and tests of this inequality hypothesis have been critiqued on methodological grounds (Kondo et al., 2009; Lynch et al., 2004; Macinko et al., 2003; Subramanian and Kawachi, 2004; Wagstaff and van Doorslaer, 2000), a formal meta-analysis did find significant associations between inequality and mortality or self-rated health in high-income countries (Kondo et al., 2009). Thus, relative position in a socio-economic hierarchy and the steepness of such hierarchies seem to matter for health.

The most cited mechanism for hierarchy-health associations is that hierarchies cause psychosocial stress, which in turn leads to poorer health outcomes (Chen and Miller, 2013; Pickett and Wilkinson, 2015). Chronic stress leads to altered hypothalamic-pituitary-adrenal (HPA) axis function, including chronically elevated cortisol levels. Increased cortisol can cause neural atrophy, cardiovascular damage, obesity, or immunosuppression, all resulting in increased susceptibility to chronic and infectious disease (Aiello et al., 2018; Garcia et al., 2017; Kunz-Ebrecht et al., 2004; Quon and McGrath, 2014; Sapolsky, 2004). In addition, submission in status competition and learned helplessness are associated with depression in humans and other primates (Hagen, 2011; Nesse, 2000; Stieglitz et al., 2014). Furthermore, experimental studies in nonhuman primates show that dominance rank affects gene expression and immune function (Snyder-Mackler et al., 2016; Tung et al., 2012). Related results in humans show that early life experiences and other forms of social stress are also associated with increases in inflammation and blunted immunological responses to cortisol (Aiello et al., 2018; Miller et al., 2014, 2011, 2009).

But why are hierarchies stressful or otherwise detrimental to health? Here we examine hierarchy-health associations from an evolutionary-medicine perspective, which suggests that many detrimental health outcomes may result from *tradeoffs* as short-term fitness gains are prioritized over long-term health, or from evolutionary *mismatch* as our bodies struggle to deal with conditions never encountered by our ancestors (Eaton et al., 2002; Gluckman et al., 2016; Nesse and Williams, 1994; Wells et al., 2017). First, given the consistent fitness benefits of high status regardless of how status is achieved in a given society (Stulp et al., 2016; Von Rueden and Jaeggi, 2016), and given that fitness is always relative, humans arguably have evolved motivations for status-striving that are independent of one’s absolute access to resources (Johnson et al., 2012). Status-striving activates the stress response, and not just for low-rankers: depending on how rank is achieved and maintained, high-or low-ranking individuals may be more stressed (Abbott et al., 2003; Sapolsky, 2005). Crucial to who is stressed is the availability of social support, which can be as or even more important for health and fitness as rank per se (Sapolsky, 2005; Snyder-Mackler et al., 2020). Other factors primarily impact low-ranking individuals: in many primate (and some human) societies, subordinates are regularly subjected to aggression and intimidation by higher-ranking individuals (Silk, 2003), resulting in the lack of control and learned helplessness that often cause depression (Sapolsky, 2005, 2004). Greater inequality, i.e. steeper hierarchies entail more skewed payoff distributions, favoring more intense competition and risk-taking, especially among low-ranking individuals; this is argued to explain the persistent association between income inequality and homicide rates, as most homicides result from escalated contests over status (Daly and Wilson, 1997, 1988). If skewed pay-off distributions and oppression of low-rankers favor “faster” life-history strategies (*sensu* Wells et al., 2017), this could also explain hierarchy-health associations via present-oriented decision-making at the expense of long-term health (Daly and Wilson, 1997; Griskevicius et al., 2011; Pepper and Nettle, 2014). Conversely, social adversity may affect health via developmental constraints (Lea et al., 2017; Snyder-Mackler et al., 2020). All of this should be true even when hierarchies are based on prestige, rather than dominance, since prestige-based hierarchies still correlate with social support, insulation against shocks, influence, sense of control, and access to mates (Gurven et al., 2000; Sugiyama and Sugiyama, 2003; von Rueden et al., 2014). Thus, stress and negative health consequences can result from perpetual status-striving, the distribution of social support, from lack of control and learned helplessness, from intensified competition especially among low-ranking individuals, and perhaps from generally “faster” life histories or developmental constraints.

In summary, the argument for why and how being low in a hierarchy negatively influence health is that humans, much like other primates, are sensitive to their relative rank and the distribution of fitness outcomes, and that we adjust our behaviors and physiological responses accordingly. Several open questions remain, however. First, the inequality hypothesis remains hotly debated, and testing it requires careful statistical methods. Second, it remains unclear to what extent the observed health consequences of relative status and possibly inequality in high-income countries represent (i) tradeoffs of potentially adaptive responses to lower relative rank and/or to inequality, or (ii) are caused by evolutionary mismatch, i.e. novel conditions that cause maladaptive outcomes. If the health consequences stem from tradeoffs to adaptive responses, e.g. people taking more risks and making more present-oriented choices at the expense of long-term health (Daly and Wilson, 1997; Griskevicius et al., 2011; Pepper and Nettle, 2014), then hierarchy should be associated with health in any population. However, if the impacts of status and inequality are caused by evolutionary mismatch, as our urban, industrialized societies combine limited upward mobility, structural violence – often based on racism – and lack of kin support with obesogenic diets and chronic inflammation (Gravlee, 2009; Sapolsky, 2004), then we would not necessarily expect hierarchy-health relationships in all societies. In particular, small-scale societies practicing traditional subsistence lifestyles (henceforth: “traditional societies”) are an important test case as they are ethnically homogenous, generally have more informal, egalitarian hierarchies (Borgerhoff Mulder et al., 2009; Kaplan et al., 2009; Mattison et al., 2016) and the major sources of morbidity and mortality are from infectious rather than chronic disease (Eaton et al., 1988; Gurven et al., 2007, 2016; Gurven and Kaplan, 2007; Kaplan et al., 2017; Pontzer et al., 2018). Further, many traditional societies have immune systems that are well calibrated by frequent exposure to pathogens and microbiota, and predominantly experience acute responses to infections (Blackwell et al., 2016a; McDade, 2005), unlike the chronic low-grade inflammation that links stress to hypertension, cardiovascular disease, and depression in high-income countries (Gurven et al., 2008). Lastly, competition (for mates, resources, etc.) in such societies is usually fairly local, meaning that the scale at which relative rank and inequality should be measured is more obvious than in large-scale modern societies with television and social media, where people are simultaneously part of many hierarchies. Thus, traditional societies present an important test case for the universality of hierarchy-health associations, i.e. whether they are likely caused by tradeoffs expected in any society or by evolutionary mismatch in modern, industrialized populations.

Few studies have examined associations between rank or inequality and health in traditional societies. Among Dominican farmers, socio-economic indicators were unrelated to cortisol levels whereas local influence was associated with lower cortisol (Decker, 2000). Among egalitarian Garisakang horticulturalists in Papua New Guinea, higher income coming from greater market exposure was associated with higher cortisol, whereas other SES measures as well as status based on locally relevant measures were not (Konečná and Urlacher, 2017). Among Tsimane forager-horticulturalists in Bolivia, the population studied here, traditional forms of status generally supported a status-health gradient, but studies on income or wealth showed mixed results. In a sample of four communities, politically more influential men had lower cortisol and a lower incidence of respiratory infection, though there were also many null results, and higher income was associated with higher cortisol (von Rueden et al., 2014). Across 13 Tsimane villages, relative wealth was associated with better self-reported health (Undurraga et al., 2010); however, average self-reported health was lower in wealthier villages. In a larger sample of Tsimane villages, relative income associated with lower (higher) BMI among individuals with smaller (larger) support networks (Brabec et al., 2007). While results are mixed, there is some converging evidence that suggests market integration generates psychosocial stress in traditional societies.

In terms of the relationship between inequality and health, studies among Tsimane have also shown mixed results. One study found no association between income inequality and body fat (Godoy et al., 2005), but income inequality was associated with more negative emotions (Godoy et al., 2006). Greater village wealth inequality did not associate with self-reported health in one study (Undurraga et al., 2010) but associated with *better* self-reported health and *lower* self-reported stress in another, controlling for individual and village wealth level (Undurraga et al., 2016). Overall, these results provide mixed evidence for associations between inequality and health.

Here we test for links between hierarchy and health among the Tsimane, expanding upon previous studies in several ways. First, we simultaneously assess the effects of relative resource access and resource inequality, controlling for absolute resource access (measured as relative wealth, wealth inequality, and absolute wealth, respectively), thus addressing major methodological critiques of the hierarchy-health hypothesis (Kondo et al., 2009; Lynch et al., 2004; Macinko et al., 2003). Second, while previous studies have mostly relied on just one or two health outcomes, we include thirteen different dependent variables (Table 1) capturing psychosocial factors and various health outcomes, including infectious disease morbidity. Third, we explicitly test whether psychosocial variables mediate links between wealth and health, as predicted if the adverse health effects of hierarchy occur through stress. Fourth, we greatly increase the sample size relative to previous studies with inequality measured in 35 communities and wealth in 1,005 households, representing approximately one third of the adult Tsimane population. Thus, our study represents the most comprehensive test of hierarchy-health associations in a traditional society.

We test the following predictions. Under the tradeoff hypothesis, which posits that human behavior and physiology are adjusted according to one’s position in a hierarchy, potentially resulting in detrimental health effects even where status hierarchy is informal and wealth differences relatively minimal, we predict that:

> *P1: Higher relative wealth is associated with better psychosocial and health outcomes, independent of absolute wealth*
>
> *P2a: Greater wealth inequality is associated with worse psychosocial and health outcomes, and P2b: this should hold especially for low-rankers*
>
> *P3: Psychosocial variables mediate wealth-health links found under P1 and P2*

These predictions are based on the assumption that household wealth is a good measure of one’s position in a status hierarchy; this assumption is commonly made in studies of industrialized nations (Pickett and Wilkinson, 2015). While social status among the Tsimane is multidimensional (von Rueden et al., 2008), household wealth indeed correlates with subjective status in the present sample (see Methods).

If, on the other hand, hierarchy-health associations in high-income countries are predominantly caused by evolutionary mismatch stemming from the combination of a) rigid socio-economic hierarchies, and b) conditions favoring chronic disease (e.g. obesogenic diets, sedentary lifestyle, chronic inflammation) and psychosocial stress (e.g. residential isolation from kin), we predict that among the Tsimane:

> *P4a: Associations between relative wealth or inequality and health will be absent or inconsistent and not necessarily mediated by psychosocial stress*
>
> *P4b: Associations between relative wealth or inequality and health will only be found for outcomes associated with chronic disease risk (e.g. BMI, blood pressure), but not necessarily with acute illness such as infectious disease*

These predictions are based on the assumptions that, while material wealth is a novel source of status among the Tsimane, it has not yet led to the kind of rigid socio-economic hierarchies typical of industrialized populations; and that the relationship between hierarchy and BMI or blood pressure is relatively linear, i.e. that these measures will respond to hierarchy even at levels that are not yet associated with chronic disease. Table 1 gives an overview of all variables and their predicted associations with wealth and inequality. In addition to the predictions listed in the table, testing P2b involves exploring interactions between inequality and relative wealth, and testing P3 involves adding psychosocial variables as covariates in models of health outcomes (see Methods).

**Table 1:** Overview of study variables and descriptive statistics. The last column helps clarify the direction of predicted associations; e.g. P1 predicts better outcomes for richer people, hence negative associations with depression, conflicts, etc. and positive associations with labor partners and self-rated health

| Variable | N | Median | SD | Range | Higher is |
| --- | --- | --- | --- | --- | --- |
| **Outcomes: Psychosocial** |  |  |  |  |  |
| Depression [sum score] | 663 | 40 | 7.14 | 23-62 | Worse |
| Conflicts [count] | 393 | 2 | 0.69 | 0-4 | Worse |
| Labor Partners [count] | 392 | 2 | 1.95 | 1-13 | Better |
| Non-social Problems [count] | 390 | 3 | 1.16 | 0-7 | Worse |
| Urinary Cortisol [pg/ml] | 828 | 154,02 | 149,526 | 93-851,308 | Worse |
| **Outcomes: Health** |  |  |  |  |  |
| BMI [Z]<sup>a</sup> | 9378 | 0.08 | 1.1 | -3.95-3.99 | ? |
| Systolic Blood Pressure [mmHg] | 1611 | 110 | 12.55 | 60-178 | Worse |
| Diastolic Blood Pressure [mmHg] | 1611 | 67 | 9.33 | 35-110 | Worse |
| Self-rated Health [scale] | 850 | 4 | 0.49 | 1-5 | Better |
| Total Morbidity [count]<sup>b</sup> | 3140 | 1 | 0.99 | 0-5 | Worse |
| Infections/parasites [yes/no]<sup>b</sup> | 3140 | 19.1% yes |  |  | Worse |
| Respiratory disease [yes/no]<sup>b</sup> | 3140 | 32.3% yes |  |  | Worse |
| Gastrointestinal [yes/no]<sup>b</sup> | 3140 | 38.8% yes |  |  | Worse |
| **Predictors** |  |  |  |  |  |
| Age [years] | 4546 | 16.97 | 17.56 | 0-89.7 |  |
| Sex [0=female, 1=male] | 4572 | 48.3% female |  |  |  |
| Household wealth [Bs] | 1003 | 5,476 | 6,125 | 211-87,772 |  |
| Community Size [count of households sampled] | 35 | 16 | 18.98 | 9-81 |  |
| Distance to market town [km] | 34 | 34.27 | 41.36 | 4.71-135.83 |  |
| Mean Community Wealth [Bs] | 35 | 8,379 | 1,995 | 4,447-12,397 |  |
| Community Wealth inequality [Gini] | 35 | 0.27 | 0.06 | 0.15-0.43 |  |

<sup>a</sup> Whether higher or lower BMI is better is a bit ambiguous: in high-income countries higher BMI is associated with worse health, lower status and greater inequality, whereas in low-income countries the reverse may be true.
<sup>b</sup> see Supplementary Table S1 for an overview of the most common morbidities by category

### Study Population

The Tsimane are a population of ∼16,000 indigenous Amerindians living in > 90 communities at the edge of the Amazon basin in lowland Bolivia. Tsimane communities consist of dispersed household clusters tied together by networks of kinship, cooperative production and consumption (Hooper et al., 2015; Jaeggi et al., 2016) as well as usually a school and soccer field. Community meetings convene to discuss and resolve important matters, including conflicts within the community. As such, we treat the community as the salient scale of status competition (Alami et al., 2020; von Rueden et al., 2018, 2008, 2019, 2014), and calculated relative wealth and inequality at this level.

The Tsimane remained relatively isolated from the larger Bolivian economy until the 1970’s and still widely practice traditional subsistence (swidden horticulture, hunting, and fishing), which contributes > 90% of their calories (Gurven et al., 2017; Kraft et al., 2018). Over the past few decades, wage labor opportunities with loggers or ranchers and produce sales in the local market town of San Borja have been increasing, as have formal schooling, Spanish fluency, and access to modern amenities such as electricity and health care. Strong, quantifiable gradients of modernization thus exist in this population (see Fig. 1).

In terms of morbidity and mortality, the Tsimane are characterized by high infectious disease burden, with respiratory infections as the leading cause of death at all ages (Gurven et al., 2007). Additionally, parasites, such as helminths and giardia are highly prevalent (Blackwell et al., 2013). This results in frequent, acute immune responses (Blackwell et al., 2016a) but a low incidence of chronic conditions such as hypertension or atherosclerosis (Gurven et al., 2012, 2016, 2009; Kaplan et al., 2017).

## Results

Wealth varied considerably by age (Fig. 1A), hence we used an age-corrected measure of relative wealth (see Methods) that reflects one’s trajectory along this age gradient. At the high end of the wealth distribution (Fig. 1B), much of the variation was driven by livestock, especially cattle, which were introduced to the region by ranchers but are only owned by a small minority of Tsimane. There was substantial variation in mean wealth and wealth inequality among the study communities (Fig. 1C-D). Mean wealth was generally lowest in communities located in the interior forest (Fig 1C, bottom right), which is remote and inaccessible by road for much of the year (due to washed out bridges), and those downriver from San Borja (Fig 1C, top), which experience frequent flooding and are within or adjacent to a protected bioreserve that limits resource extraction to residents. We operationalized inequality by calculating a village-level Gini coefficient for wealth (see Methods). Wealth inequality was generally lower in communities farther from the market towns of San Borja and Yucumo, where Tsimane can sell produce and purchase market goods, though some villages near towns also show low inequality (Fig 1D).

**Fig. 1:**
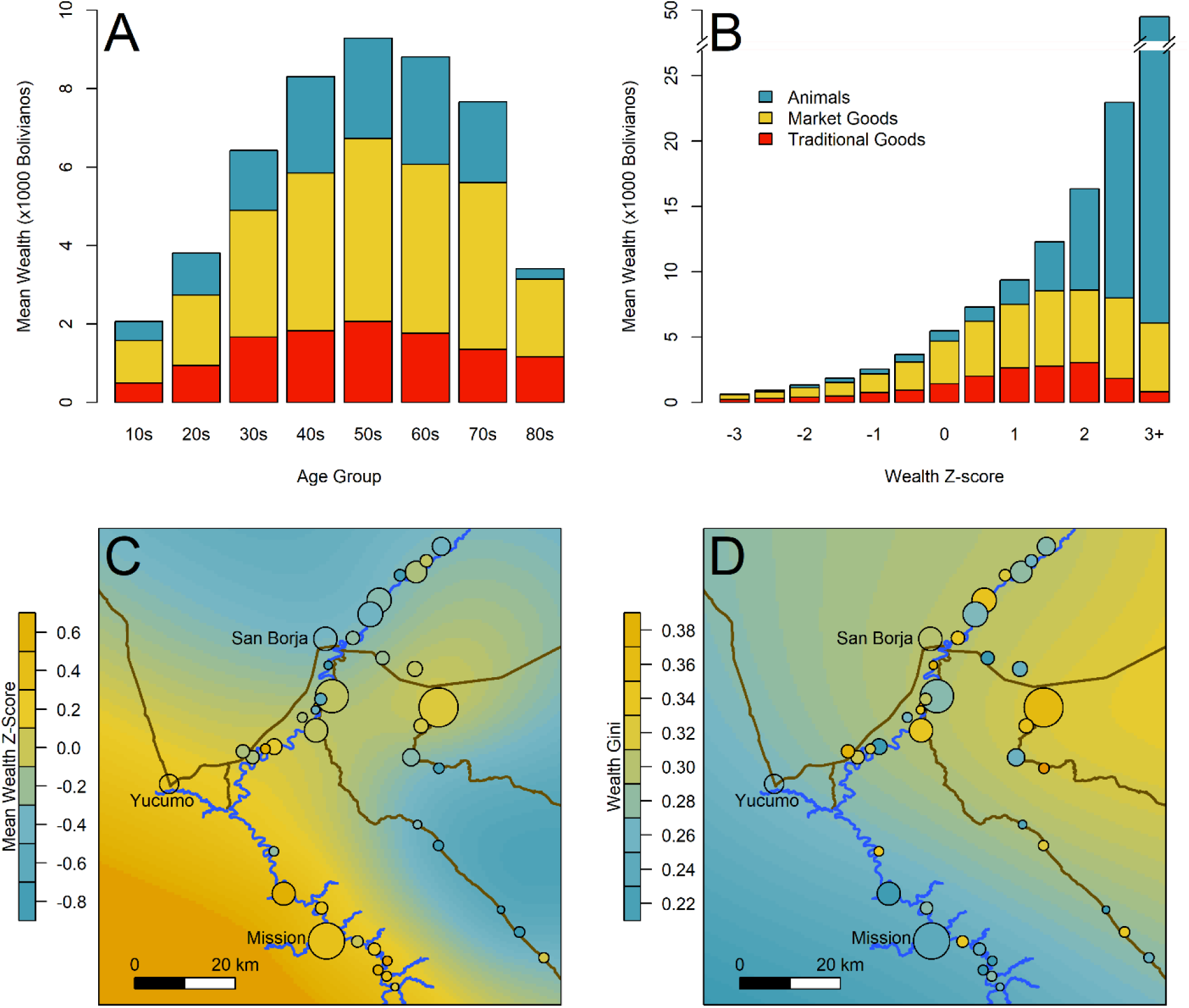
A) Mean wealth by age of household head. B) Mean wealth by population-level wealth Z-score. C) Map of study communities (n = 35) and mean wealth at the community level. D) Map of community-level wealth inequality. Note: A and B use raw wealth, while C and D are based on age-corrected values. Heat maps in C and D give a rough sense of the distribution, while colored circles indicate actual values for each community; circle size indicates the number of sampled households (range = 9–81). Yucumo and San Borja are local market towns inhabited by non-Tsimane, Mission is the site of a Catholic mission and the largest Tsimane settlement.

To examine the effects of household wealth and community wealth inequality on psychosocial or health outcomes, we ran Bayesian multilevel models with appropriate controls and random effects at the individual, household, and community level (see Methods). Importantly, by including mean wealth at the community level, we are able to disentangle *absolute* and *relative* wealth effects. Specifically, household wealth Z-scores were centered within each community (see Methods), thus reflecting relative wealth within a community (van de Pol and Wright, 2009). Absolute wealth associations were calculated as the combined posterior effects of community-level mean wealth and within-community wealth Z-scores. Results reported in the main text focus mostly on relative wealth and inequality as the pertinent tests of our predictions, and are reported as standardized coefficients (β) or odds ratios (OR), both represented by the posterior mean, as well as the proportion of the posterior on one side of zero (P_>0_ or P_< 0_), i.e. the probability of a positive or negative association. Means and credible intervals for all parameters are reported in supplementary tables S2-S14. In most models, the predictors and random effects jointly explained about 20–40% of the variance in the data (R^2^ range: 0.16–0.91, see Tables S2-S14).

There were some beneficial associations between relative household wealth and psychosocial or health outcomes consistent with P1 (Fig. 2). In terms of psychosocial outcomes, greater relative wealth was associated with having more labor partners (β = 0.16, P_>0_ = 1.00). Associations with fewer depressive symptoms (β = –0.04, P_< 0_ = 0.85), non-social problems (i.e. self-reported concerns over food insecurity, debt, and illness; β = –0.06, P_< 0_ = 0.87), and urinary cortisol (β = –0.02, P_< 0_ = 0.76) were weaker and more uncertain. There was no support for an association with social conflicts. In terms of health, relative wealth was associated with lower total morbidity (β = –0.03, P_< 0_ = 0.92), and lower odds of respiratory illness (OR = 0.88, P_< 1_ = 0.98). The association with self-rated health (β = 0.03, P_>0_ = 0.78) was highly uncertain but in the expected direction. The only detrimental association was with gastrointestinal illness, which was more likely in relatively wealthier people (OR = 1.07, P_>1_ = 0.93). The were no or very weak associations between relative wealth and BMI, systolic and diastolic blood pressure, and other infections. However, absolute wealth was strongly associated with lower systolic (β = –0.32, P_< 0_ = 0.99) and diastolic (β = –0.38, P_< 0_ = 0.98) blood pressure (Figure S1, Tables S7 & S8); people with higher absolute wealth also had more labor partners (β = 0.33, P_>0_ = 0.88). Other than blood pressure and labor partners, absolute wealth was not associated with any outcomes, reaffirming the notion that relative rank matters more than absolute resource access.

To examine whether psychosocial variables mediated associations between relative wealth and health (P3), we included psychosocial variables (depression, non-social problems, and cortisol levels) as covariates in models of blood pressure, self-rated health and morbidities (Tables S8-S14). This did not change the strength or certainty of wealth-health associations in any meaningful way, despite several psychosocial variables being themselves strongly associated with the health outcomes and sometimes substantially improving goodness of fit of these models. Specifically, depression was strongly associated with worse self-rated health (β = –0.24, P_< 0_ = 1.00), more total morbidities (β = 0.12, P_>0_ = 1.00), and a greater risk of respiratory (OR = 1.21, P_>1_ = 0.85) and gastrointestinal illness (OR = 1.26, P_>1_ = 0.92). Having fewer non-social problems was associated with reduced total morbidity (β = –0.10, P_< 0_ = 1.00) and a reduced risk of gastrointestinal illness (OR = 0.70, P_< 1_ = 1.00); each SD increase in cortisol associated with a greatly increased risk of respiratory illness (OR = 2.61, P_>1_ = 1.00). This indicates that psychosocial variables are indeed associated with health outcomes (Stieglitz et al., 2015), but independently of wealth.

**Fig. 2:**
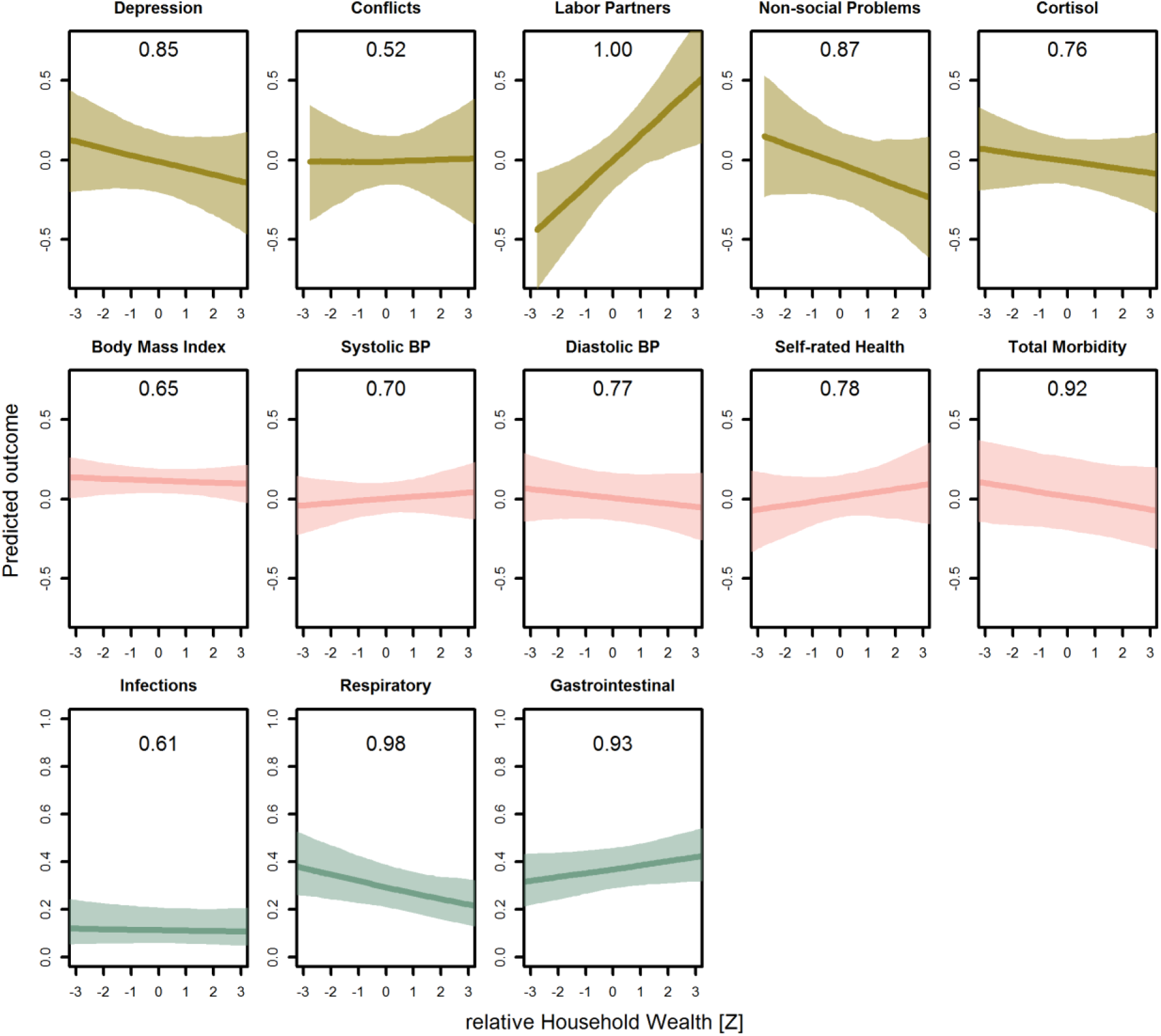
Predicted associations between relative household wealth, i.e. within-community Z score, and all psychosocial and health outcomes. Lines are posterior means and shaded areas are 95% credible intervals. Numbers in each panel represent the posterior probability, i.e. the proportion of the posterior distribution that supports an association between wealth and the outcome (P_>0_ or P_< 0_). All predictions control for age, sex, inequality, distance to market town, community size, and mean community wealth, holding all other variables at the mean, with sex = female. For the first two rows, the outcomes are measured as Z scores, the bottom row as probabilities. Rough categories of dependent variables (psychosocial, continuous health outcomes, and binary health outcomes) are distinguished by rows and colors.

For wealth inequality (Fig 3, Tables S2-S14), associations with psychosocial variables were mostly negligible, with the strongest being *fewer* non-social problems in more unequal communities (β = –0.15, P_< 0_ = 0.92). However, wealth inequality was associated with some detrimental health outcomes, consistent with P2a: greater inequality was associated with higher blood pressure (systolic: β = 0.07, P_>0_ = 0.92; diastolic: β = 0.08, P_>0_ = 0.89), worse self-rated health (β = –0.10, P_< 0_ = 0.92), and a greater likelihood of respiratory illness (OR = 1.36, P_>0_ = 0.95). Despite these harmful associations with inequality, people in more unequal communities had a strongly reduced likelihood of other infections (OR = 0.63, P_< 0_ = 0.96) and to a more uncertain degree, total morbidity (β = –0.07, P_< 0_ = 0.77). As with relative wealth, we examined whether psychosocial variables would mediate inequality-health associations (Tables S8-S14). We found that none of the inequality-health associations were mediated by psychosocial stress, contrary to P3. Similarly, the prediction that inequality should affect poorer people more was generally not supported (see Table S15, below).

**Fig. 3:**
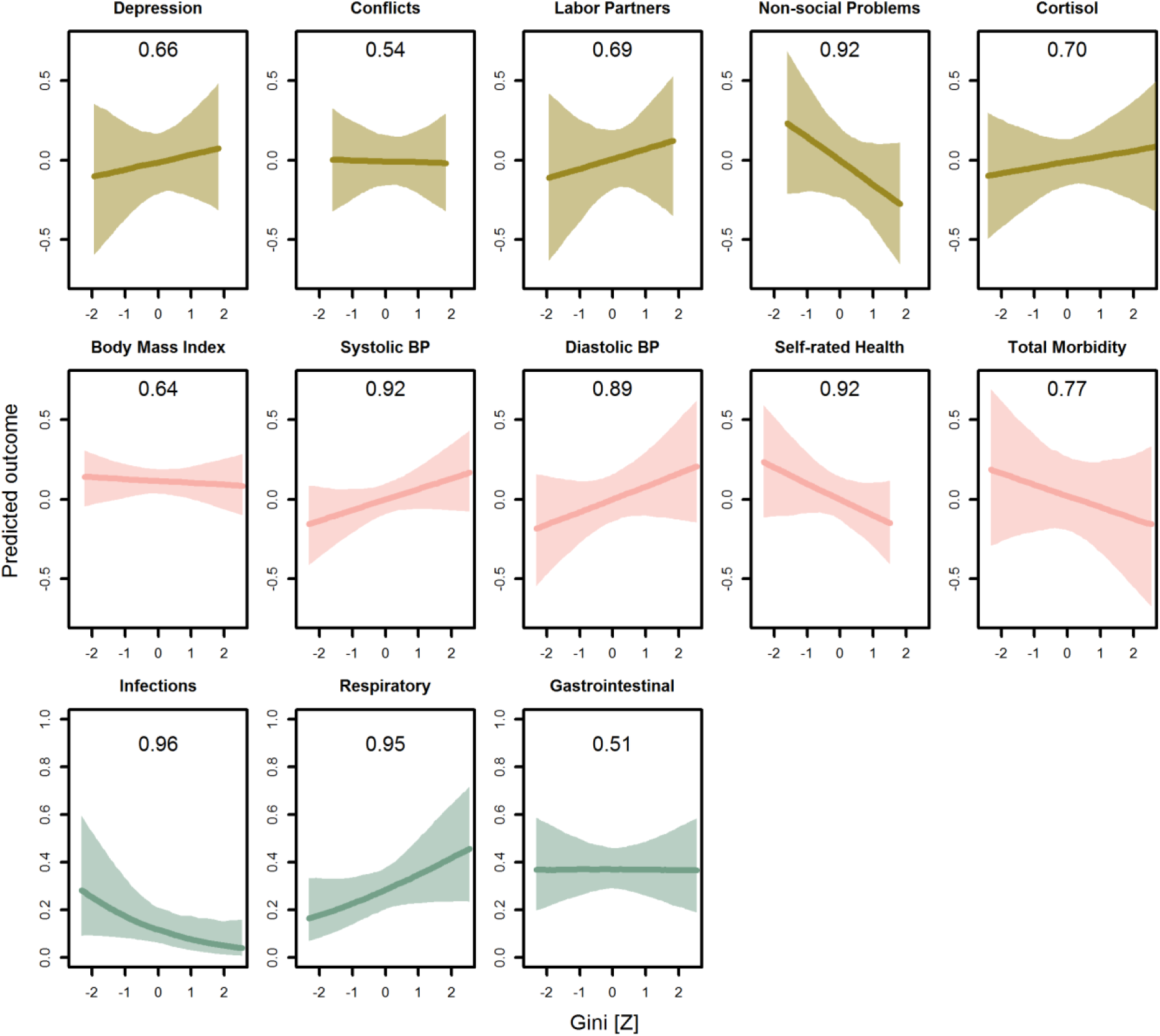
Predicted associations between wealth inequality (Gini coefficients) and all psychosocial and health outcomes. Lines are posterior means and shaded areas are 95% credible intervals. Numbers in each panel represent the posterior probability, i.e. the proportion of the posterior that supports an association between inequality and the outcome. All predictions control for age, sex, household wealth, distance to market town, community size, and mean community wealth, holding all other variables at the mean, with sex = female. For the first two rows, the outcomes are measured as Z scores, the bottom row as probabilities. Rough categories of dependent variables (psychosocial, continuous health outcomes, and binary health outcomes) are distinguished by rows and colors.

Finally, we conducted several post-hoc tests to examine whether wealth-health associations were contingent on sex, or whether relative wealth effects were contingent on levels of inequality and vice versa. For example, inequality could trigger increased stress and competitiveness only in men given a history of higher reproductive skew in males (Daly and Wilson, 1997, 1988) and inequality might affect the wealthier and poorer differently (P2b), i.e. poorer individuals may fare even worse in more unequal contexts. For this we included wealth x inequality, wealth x sex, or inequality x sex interactions. Several associations were contingent (Table S15). In particular, wealth or gini associations with depression, cortisol, diastolic blood pressure, infections, respiratory and gastrointestinal illness varied by sex, albeit not consistently. While only the labor partners and diastolic blood pressure models favored a gini x wealth interaction. Thus, inequality seems to generally affect the wealthier and poorer equally, contra P2b, while sometimes men and sometimes women are more affected by wealth or inequality.

## Discussion

We tested whether relative wealth and wealth inequality were associated with a broad range of psychosocial and health outcomes, independently of absolute wealth, in a small-scale subsistence society. We found that relative wealth within a community and community-level wealth inequality were generally more strongly associated with outcomes than absolute wealth, which clearly indicates relative position in a hierarchy matters above and beyond absolute access to resources, and reaffirms our decision to treat the community as the relevant scale of analysis. Indeed, absolute wealth was hardly associated with any outcomes (Figure S1), likely because the most common health risks affect everyone similarly within a community.

Consistent with the hypothesis that higher relative position in a socio-economic hierarchy improves outcomes and the steepness of that hierarchy worsens them (P1 and P2a), wealthier people had better psychosocial outcomes, fewer morbidities and a reduced risk of respiratory illness, while people in more unequal communities suffered from worse self-rated health and an increased risk of respiratory illness. Similarly, von Rueden et al (2014) found lower risk of respiratory infection among influential men. Thus, converging results implicate status in respiratory morbidity among the Tsimane. Given that respiratory illness is the leading cause of mortality at all ages in this population (Gurven et al., 2007), these results indicate substantial fitness costs to being outranked in a local hierarchy and to living in unequal places – for instance, the likelihood of being diagnosed with respiratory illness was predicted to differ almost 3-fold, 0.17 to 0.49, between the least and most unequal communities. These results may be especially relevant today as the Tsimane prepare for the COVID-19 pandemic (Kaplan et al., 2020), which causes higher morbidity and mortality in socio-economically disadvantaged people (Devakumar et al., 2020; Wang and Tang, 2020). Therefore, our findings support the notion that relative rank and inequality affect health – including infectious disease morbidity (Aiello et al., 2018) – even in relatively egalitarian societies. This suggests that humans universally adjust their behavior and physiology to their status and locally relevant competitive regimes (Daly and Wilson, 1997; Griskevicius et al., 2011; Pepper and Nettle, 2014) though we cannot say whether such adjustment is temporary or permanent, as conceptualized by life-history strategies or developmental constraints (Lea et al., 2017; Snyder-Mackler et al., 2020; Wells et al., 2017).

Despite some clear hierarchy-health associations, we found limited support for the hypothesis that these associations were mediated by psychosocial stress (P3). Associations between wealth and psychosocial outcomes were generally weaker and, importantly, none of the wealth-health associations were altered when including psychosocial measures, including cortisol levels, as covariates (even though cortisol was itself associated with some health outcomes). In contrast, a study of four Tsimane communities found that influential men with greater social support had lower cortisol (von Rueden et al., 2014), and higher cash income associated with higher cortisol in two traditional societies (Konečná and Urlacher, 2017; von Rueden et al., 2014). In another study of the Tsimane, higher incomes predicted lower BMIs, unless individuals had relatively more social support (Brabec et al., 2007). For traditional societies experiencing market integration, whether relative status increases, decreases, or has no effect on stress and health may depend on the status measure and its association with social support. It therefore remains unclear what mechanisms were responsible for the wealth-health associations found here, though hierarchy is known to affect immune function, and thereby infectious disease morbidity somewhat independently of HPA activity (Aiello et al., 2018; Snyder-Mackler et al., 2020, 2016).

Arguably, our results also lend some support to the hypothesis (P4) that hierarchy-health associations in high-income countries are due to, or at least exacerbated by evolutionary mismatch (Sapolsky, 2004). Specifically, wealth-health associations were somewhat inconsistent (P4a), with some associations being negligible or counter to predictions. One of the strongest associations of inequality was with blood pressure, a major contributor to chronic disease (P4b). While the vast majority of Tsimane do not suffer from clinical levels of hypertension or associated cardiovascular disease (Gurven et al., 2012; Kaplan et al., 2017), the predicted effects of inequality on blood pressure were substantial: systolic blood pressure was predicted to increase by 0.32 SD (i.e. 4.0 mmHg) and diastolic blood pressure by 0.40 SD (3.7 mmHg) in the most unequal compared to the most equal communities. In Western countries such a change in blood pressure corresponds to about a 10% change in the risk of major cardiovascular disease events (see Figure 2 in Ettehad et al., 2016). Among the Tsimane, it corresponds to 20 years (systolic) or 40 years (diastolic) of age-related increases in blood pressure (Gurven et al., 2012). As novel, obesogenic foods enter the Tsimane diet (Kraft et al., 2018), market integration increases stress (Konečná and Urlacher, 2017; von Rueden et al., 2014), and protective lifestyle factors like physical activity and helminth infections are changing (Gurven et al., 2013, 2016), people in unequal communities, especially the poor (see Figure S1), may be at greater risk of chronic disease. In this context, it is also worth noting that while the range of our village-level Gini values (0.15–0.43) was similar to that of *income* inequality among high-income countries (e.g. Denmark: 0.24, USA: 0.45), it was considerably lower than the range of *wealth* inequality (e.g. Japan: 0.55, USA: 0.81 (Nowatzki, 2012)). Thus, assuming that the associations between hierarchy and blood pressure persist across a wider range, we found support for the prediction (P4b) that hierarchy contributes to chronic disease risk.

In sum, we present the most comprehensive test of hierarchy-health associations in a traditional society to date. Thanks to more detailed data and better methods than those used in many studies in high-income countries, we also provided a stronger test of the relative wealth and inequality hypotheses than most (Kondo et al., 2009; Lynch et al., 2004; Macinko et al., 2003; Wagstaff and van Doorslaer, 2000). In support of an evolutionary argument that conceptualizes hierarchy-health effects as stemming from evolved reaction norms adjusting people’s behavior and physiology to the rank and local competitive regime they find themselves in (Daly and Wilson, 1997; Griskevicius et al., 2011; Pepper and Nettle, 2014), we found that relative wealth and inequality were strongly associated with several health outcomes. In support of the argument that most hierarchy-health effects in high-income countries are caused by evolutionary mismatch (Sapolsky, 2004), we found that inequality was associated with blood pressure even in this relatively egalitarian society, which could lead to hypertension, cardiovascular and metabolic disease as inequality further increases and/or novel foods and lifestyle factors enter the population (Gurven et al., 2012, 2016; Kaplan et al., 2017; Kraft et al., 2018). Our study thus contributes to an evolutionary approach to public health that considers evolved reaction norms and mismatch as important links between lifestyle and health (Eaton et al., 2002; Wells et al., 2017).

## Materials and Methods

### Data collection and preparation

All data were collected under the auspices of the Tsimane Health and Life History Project (THLHP) (Gurven et al., 2017) by a team of Bolivian medical professionals and Tsimane researchers.

#### Wealth and wealth inequality

Wealth data were collected in 2006/2007 and 2013/2014. Household wealth was assessed through an inventory of commonly owned items including traditional goods, i.e. items manufactured from local organic materials (e.g. canoes, bows and arrows), market goods, i.e. industrially produced items obtained through trade or purchase (e.g. bicycles, motorbikes), and livestock (e.g. pigs, cows), which were subsequently converted into their local market value in Bolivianos and summed (Fig. 1).

Objective household wealth arguably provides only an indirect measure of people’s subjective wealth and status (Norton, 2013) but these data were most widely available for this study. Furthermore, household wealth correlated significantly, albeit weakly, with subjective status (Amir et al., 2019; Woolard et al., 2019; r = 0.17, df = 147, P< 0.05) and subjective wealth rank (r = 0.29, df = 150, P< 0.001). Previous work among the Tsimane (Undurraga et al., 2016) has also shown that more visible forms of wealth, such as the household items counted here, influenced subjective health more than less visible forms of wealth, such as the size of cultivated fields. To prevent differences in age sampling between villages from affecting wealth and inequality estimates, we followed Borgerhoff Mulder et al (2009) and adjusted wealth values for the age of the head of household by fitting generalized additive models for location scale and shape (GAMLSS) to the distribution of wealth-by-age to obtain wealth-by-age zscores.

Mean wealth and wealth inequality at the community level (for communities with > = 9 households) were calculated after converting wealth Z-scores back into equivalent values in Bolivianos at age 50 (see Fig. 1). We used the Gini coefficient to measure inequality; other inequality measures (e.g. median share, 90/10 ratio) generally correlate highly (r>0.94) with Gini (Kondo et al., 2009) and were therefore not considered. In other studies, local scales of measuring inequality, such as at the community level used here, tend to produce smaller effects on health than those at larger scales, such as states or countries (Kondo et al., 2009; Wilkinson and Pickett, 2006). In the Tsimane context, it is unclear whether that will be the case given low residential mobility and concentration of work and socializing within communities. However, Tsimane will occasionally visit families in other communities and sporadically engage in market-based interaction with non-Tsimane, and comparisons with wealther neighbors can contribute to Tsimane status aspirations (Schultz, 2019). Nevertheless, as mentioned above (Study population), we consider the community to be the most relevant arena for status competition among Tsimane. Note that most studies on health effects of inequality use *income* inequality (but see Nowatzki, 2012), which is less unequally distributed than wealth. Cash income among the Tsimane during this study period was sporadic and many households may have no income in a given sampling period, which leads to overestimated Ginis. We therefore preferred wealth and wealth inequality as a more reliable measure of households’ long-term access to resources and its distribution.

#### Psychosocial and health variables

A variety of variables recorded during roughly annual examinations by THLHP physicians and anthropologists were merged onto the wealth data; all data collected within two years of the wealth data were considered. Body mass index (BMI) Z-scores were calculated using Tsimane-specific growth curves (Blackwell et al., 2016b) (http://www.github.com/adblackwell/localgrowth) as well as the total distribution of Tsimane adult BMIs, representing deviations from the local population average for a given age and sex. Diastolic and systolic blood pressure were measured by THLHP physicians during routine medical exams. Cortisol was measured in first-morning urine, corrected for specific gravity using enzyme-linked immunosorbent assays (see von Rueden et al., 2014 for details).

Depressive symptoms were measured using an adapted 18-item questionnaire (Stieglitz et al., 2014), the responses to which were summed to yield an overall depression score. The same interview also asked whether participants experienced conflicts with several kinds of social partners as well as non-social problems, such as food insecurity, illness, or debt; affirmative answers were summed to yield a composite measure of social conflicts and non-social problems, respectively. A household’s cooperation network was measured as the number of people from different households who helped in that household’s fields in a given year.

Morbidity was assessed during regular medical check-ups using the ICD-10 classification (International Classification of Disease, 10^th^ edition) and then grouped into 18 clinically meaningful categories following the Clinical Classifications System (https://www.hcupus.ahrq.gov/toolssoftware/ccs/ccsfactsheet.jsp); morbidities in any of these categories were summed to give a total morbidity score potentially ranging from 0 (no morbidities) to 18 (at least one morbidity in each category). In addition, we also examined the presence/absence of infectious and parasitic diseases (CCS 1, hereafter “infections”), diseases of the respiratory system (CCS 8, “respiratory illness”) and diseases of the digestive system (CCS 9, “gastrointestinal illness”), which represent the most common causes of morbidity and mortality in this population (Gurven et al., 2007). See Table S1 for examples of the six most common diagnoses in these three categories. Self-rated general health was measured at each check-up using a five-point scale from (“very bad” [1] to “excellent” [5]). Distance to the town of San Borja was measured as nearest route (whether by river or road) from the center of the community and provides a proxy for access to modern amenities.

### Data analysis

Prior to analysis, all variables were transformed into z-scores (in case of cortisol levels after log transformation due to their skewed distribution) such that coefficients were standardized across outcomes for comparability. All outcomes were modeled as Gaussian, except the presence/absence of specific morbidities (Bernoulli). Each analysis modeled an individual-level outcome as a function of individual-, household-, and community-level characteristics (Table 1). Thus, we fit the following base model for each outcome:

Outcome_*ijkl*_ ∼ β_0_ + (β_1_ * Sex_*j*_) + (β_2_ * Age_*j*_)+ (β_3_ * relative household wealth_*k*_) + (β_4_ * Community-level Gini_*l*_) + (β_5_ * Community-level mean wealth_*l*_) + (β_6_ * Community Size_*l*_) + (β_7_ * Distance of community to market town_*l*_) + *u_j_* + *u_k_* + *u*_l_ + *e_ijkl_*

wherein the subscripts denote measurement *i*, individual *j*, household *k*, and community *l*, respectively. β_0_ is the intercept, all other β’s are slopes, *u*’s are random intercepts, and *e* is the residual error (not available for Bernoulli responses). Variance inflation factors indicated virtually no collinearity among predictors (all VIFs < 3). After fitting the models, the β for absolute wealth was calculated as the sum of the posterior samples for β_3_ and β_5_.

In order to test whether potential wealth-health associations were mediated by psychosocial stress we re-ran all health models (blood pressure, self-rated health, total morbidity, infections, respiratory and gastrointestinal illness) with pertinent psychosocial variables as covariates and checked whether this changed the associations with wealth and inequality. In particular, we included depression, non-social problems and cortisol levels as covariates as they had the greatest overlap with the health samples. Furthermore, PCAs revealed that depression was capturing up to 90% of the variation in psychosocial stress. Because of missing values in the psychosocial covariates, we used Bayesian imputation while fitting these models, which makes no additional assumptions on missingness compared to complete-case analysis, preserves uncertainty and leads to less biased estimates (McElreath, 2016). In addition, we also ran a series of exploratory analyses in which we added interaction terms. We reported interactions if they improved the expected log pointwise predictive density [ELPD] in a stepwise forward selection procedure.

We used Bayesian multilevel models fit with the *brms* package v. 2.8.3. (Bürkner, 2017) in R 3.5.2. for all analyses. All models used regularizing priors (fixed effects: normal, mean = 0, SD = 1; random effects: halfcauchy, location = 0, scale = 2) which imposes conservatism on parameter estimates and reduces the risk of inferential errors (Gelman et al., 2013; McElreath, 2016). All models converged well as assessed by inspecting trace plots and standard diagnostics (all Rhat = < 1.01). Bayesian models produce a posterior distribution of parameter estimates that can be summarized in various ways (McElreath, 2016). Here we plot predicted associations between wealth/inequality and all outcomes (Figs. 1&2, S1). We also report mean standardized regression coefficients β or odds ratios [OR] and the proportion of the posterior supporting a positive or negative association [P_>0_ or P_< 0_ for β’s, P_>1_ or P_< 1_ for OR’s]. Full model summaries with means and 95% CIs, as well as (conditional) Bayesian *R^2^* (Gelman et al., 2019) as a goodness of fit measure are given in the supplementary materials (Tables S2-S14). All data and R code will be made available at https://github.com/adrianjaeggi/tsimanewealthandhealth.

## Data Availability

All data and R code will be made available at https://github.com/adrianjaeggi/tsimanewealthandhealth

## Acknowledgements

We thank the Tsimane for their generous participation and years of collaboration, and THLHP personnel for their herculean efforts and dedication in data collection. We also thank the Santa Fe Institute’s working group on wealth inequality in small-scale societies, led by Monique Borgerhoff Mulder and Sam Bowles, for stimulating discussions.

## Footnotes

1 We use status or rank interchangeably to refer to one’s position in a hierarchy. In the present study, we focus on household wealth because it was most widely available, but show that it correlates with other measures of status (see Methods)

## Supplementary material

**Figure S1:**
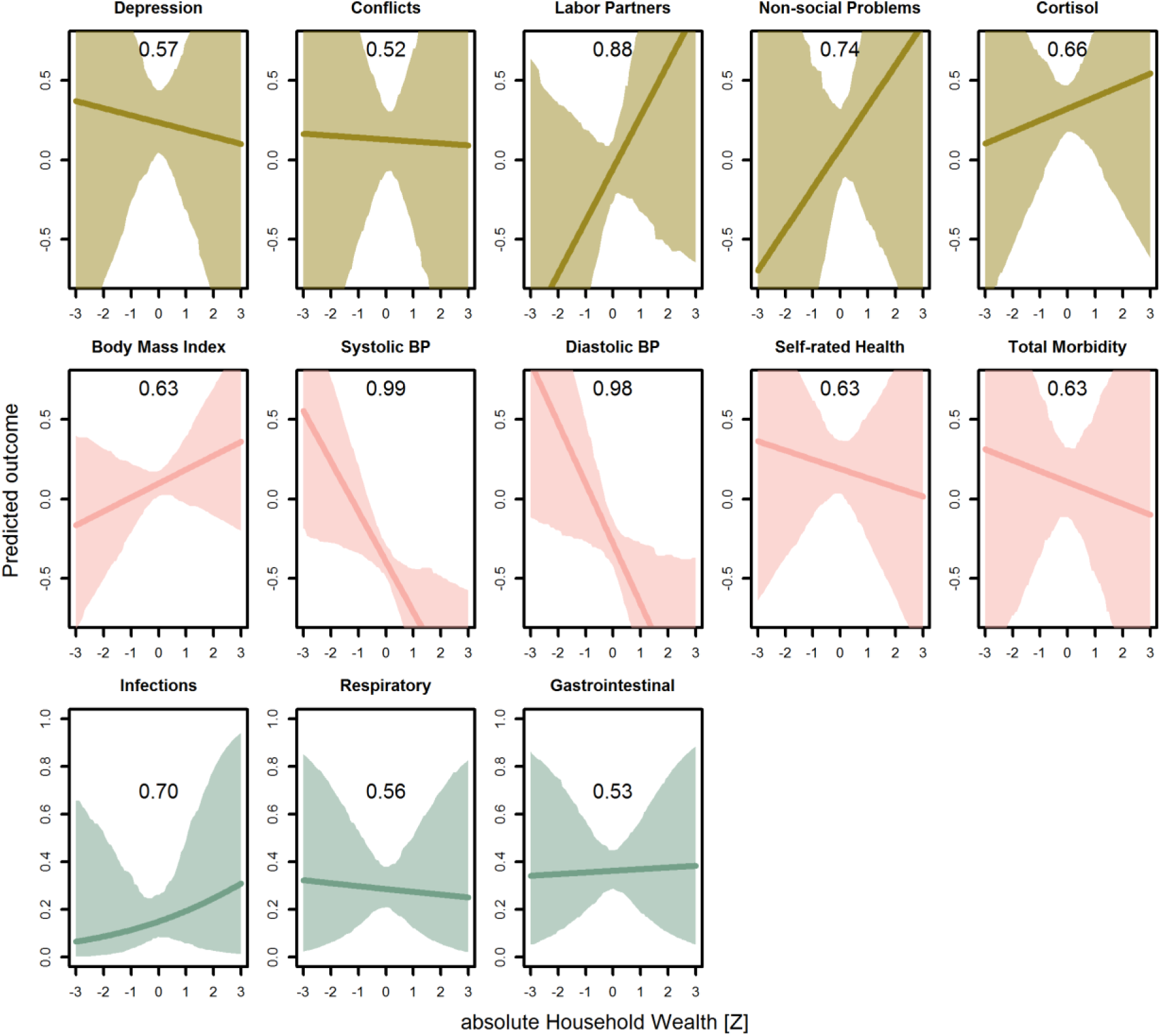
Predicted associations between absolute wealth and all outcomes. Lines are posterior means and shaded areas are 95% credible intervals. Numbers in each panel represent the posterior probability, i.e. the proportion of the posterior that supports an association between inequality and the outcome. All predictions hold all other variables at the mean, with sex = female. For the first two rows, the outcomes are measured as Z scores, the bottom row as probabilities. Rough categories of dependent variables (psychosocial, continuous health outcomes, and binary health outcomes) are distinguished by rows and colors.

**Table S1:** Overview of most common morbidities. Three of the most common Clinical Classification Systems categories (CCS, number in parentheses) and the 6 most prevalent diagnoses within each category (in decreasing order down rows, ICD-10 codes in parentheses). Musculoskeletal conditions (CCS 13) were also common but not analyzed independently

| **Infectious and parasitic diseases (CCS 1)** | **Diseases of the respiratory system (CCS 8)** | **Diseases of the digestive system (CCS 9)** |
| --- | --- | --- |
| Pediculosis due to *Pediculus humanus capitis* (B85.0) | Acute nasopharyngitis [common cold] (J00) | Intestinal helminthiasis (B82.0) |
| Tinea unguium (B35.1) | Acute streptococcal tonsillitis; unspecified (J03.00) | Infectious gastroenteritis and colitis (A09) |
| Candidiasis of vulva and vagina (B37.3) | Streptococcal pharyngitis (J02.0) | Dyspepsia (K30) |
| Pediculosis; unspecified (B85.2) | Acute upper respiratory infection; unspecified (J06.9) | Gastro-esophageal reflux disease with esophagitis (K21.0) |
| Superficial mycosis; unspecified (B36.9) | Acute bronchitis due to *Mycoplasma pneumoniae* (J20.0) | Giardiasis [lamblia] (A07.1) |
| Necatoriasis (B76.1) | Bronchopneumonia; unspecified organism (J18.0) | Gastritis; unspecified, without bleeding (K29.70) |

**Table S2:**
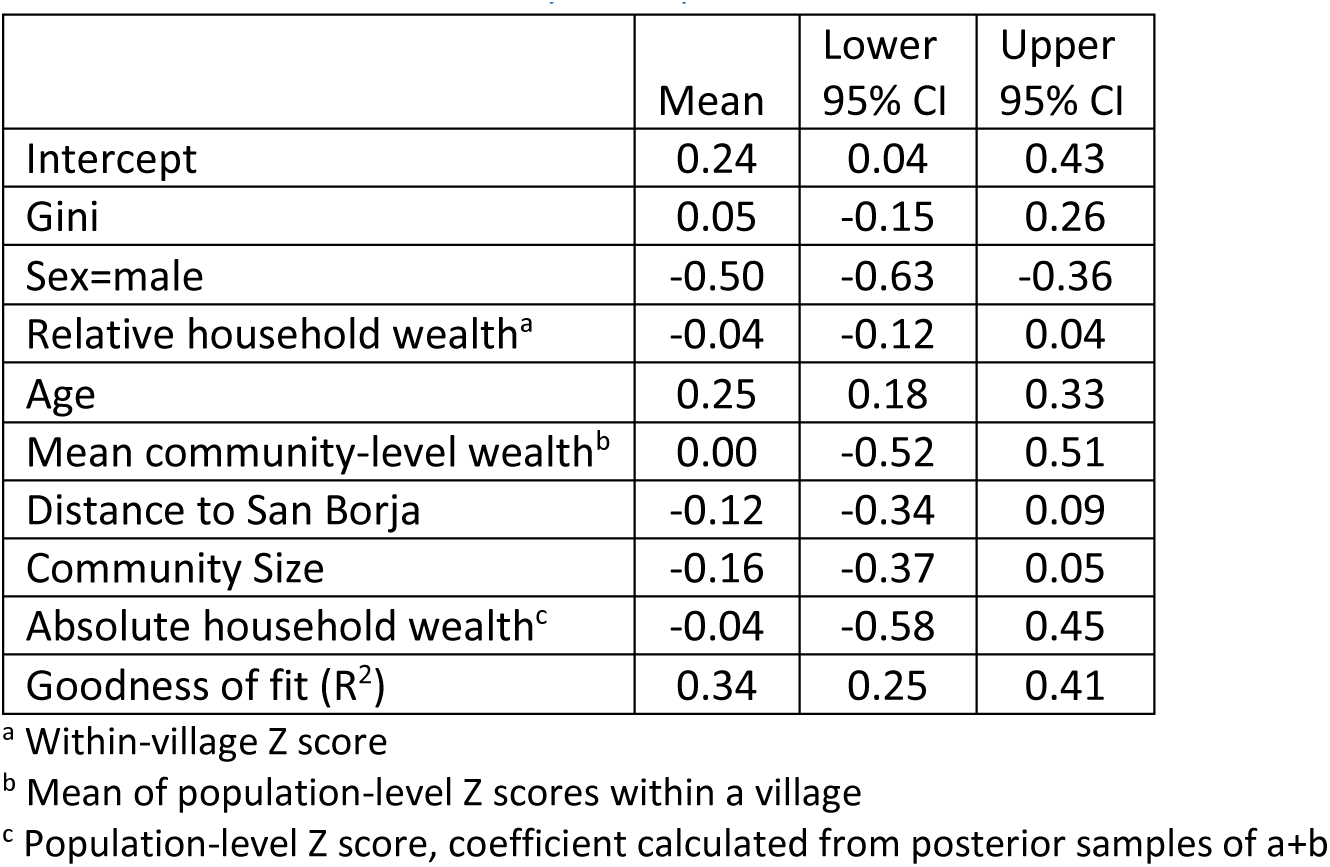
Model summary – Depression.

|  | Mean | Lower 95% CI | Upper 95% CI |
| --- | --- | --- | --- |
| Intercept | 0.24 | 0.04 | 0.43 |
| Gini | 0.05 | -0.15 | 0.26 |
| Sex=male | -0.50 | -0.63 | -0.36 |
| Relative household wealth<sup>a</sup> | -0.04 | -0.12 | 0.04 |
| Age | 0.25 | 0.18 | 0.33 |
| Mean community-level wealth<sup>b</sup> | 0.00 | -0.52 | 0.51 |
| Distance to San Borja | -0.12 | -0.34 | 0.09 |
| Community Size | -0.16 | -0.37 | 0.05 |
| Absolute household wealth<sup>c</sup> | -0.04 | -0.58 | 0.45 |
| Goodness of fit (R<sup>2</sup>) | 0.34 | 0.25 | 0.41 |

<sup>a</sup> Within-village Z score
<sup>b</sup> Mean of population-level Z scores within a village
<sup>c</sup> Population-level Z score, coefficient calculated from posterior samples of a+b

**Table S3:** Model summary – Social conflicts.

|  | Mean | Lower 95% CI | Upper 95% CI |
| --- | --- | --- | --- |
| Intercept | 0.13 | -0.07 | 0.33 |
| Gini | -0.01 | -0.17 | 0.15 |
| Sex=male | -0.08 | -0.25 | 0.11 |
| Relative household wealth<sup>a</sup> | 0.00 | -0.12 | 0.11 |
| Age | 0.18 | 0.07 | 0.30 |
| Mean community-level wealth<sup>b</sup> | -0.02 | -0.57 | 0.60 |
| Distance to San Borja | 0.20 | -0.02 | 0.42 |
| Community Size | 0.02 | -0.15 | 0.19 |
| Absolute household wealth<sup>c</sup> | -0.01 | -0.58 | 0.63 |
| Goodness of fit (R<sup>2</sup>) | 0.28 | 0.15 | 0.40 |

<sup>a</sup> Within-village Z score
<sup>b</sup> Mean of population-level Z scores within a village
<sup>c</sup> Population-level Z score, coefficient calculated from posterior samples of a+b

**Table S4:** Model summary – Labor partners.

|  | Mean | Lower<br>95% CI | Upper<br>95% CI |
| --- | --- | --- | --- |
| Intercept | -0.05 | -0.26 | 0.14 |
| Gini | 0.06 | -0.19 | 0.30 |
| Sex=male | 0.00 | -0.07 | 0.08 |
| Relative household wealth<sup>a</sup> | 0.16 | 0.05 | 0.28 |
| Age | -0.03 | -0.12 | 0.04 |
| Mean community-level wealth<sup>b</sup> | 0.17 | -0.37 | 0.75 |
| Distance to San Borja | 0.07 | -0.15 | 0.29 |
| Community Size | 0.10 | -0.07 | 0.29 |
| Absolute household wealth<sup>c</sup> | 0.33 | -0.16 | 0.97 |
| Goodness of fit (R<sup>2</sup>) | 0.91 | 0.89 | 0.92 |

<sup>a</sup> Within-village Z score
<sup>b</sup> Mean of population-level Z scores within a village
<sup>c</sup> Population-level Z score, coefficient calculated from posterior samples of a+b

**Table S5:** Model summary – Non-social problems.

|  | Mean | Lower<br>95% CI | Upper<br>95% CI |
| --- | --- | --- | --- |
| Intercept | 0.08 | -0.17 | 0.32 |
| Gini | -0.15 | -0.36 | 0.06 |
| Sex=male | -0.16 | -0.33 | 0.02 |
| Relative household wealth<sup>a</sup> | -0.06 | -0.18 | 0.05 |
| Age | 0.00 | -0.11 | 0.11 |
| Mean community-level wealth<sup>b</sup> | 0.32 | -0.39 | 1.04 |
| Distance to San Borja | 0.07 | -0.19 | 0.33 |
| Community Size | -0.16 | -0.37 | 0.05 |
| Absolute household wealth<sup>c</sup> | 0.26 | -0.50 | 0.93 |
| Goodness of fit (R<sup>2</sup>) | 0.36 | 0.24 | 0.47 |

<sup>a</sup> Within-village Z score
<sup>b</sup> Mean of population-level Z scores within a village
<sup>c</sup> Population-level Z score, coefficient calculated from posterior samples of a+b

**Table S6:** Model summary – Cortisol.

|  | Mean | Lower<br>95% CI | Upper<br>95% CI |
| --- | --- | --- | --- |
| Intercept | 0.32 | 0.17 | 0.47 |
| Gini | 0.04 | -0.11 | 0.18 |
| Sex=male | -0.79 | -0.92 | -0.67 |
| Relative household wealth<sup>a</sup> | -0.02 | -0.09 | 0.04 |
| Age | -0.05 | -0.12 | 0.02 |
| Mean community-level wealth<sup>b</sup> | 0.10 | -0.26 | 0.47 |
| Distance to San Borja | -0.08 | -0.22 | 0.05 |
| Community Size | -0.08 | -0.24 | 0.07 |
| Absolute household wealth<sup>c</sup> | 0.07 | -0.31 | 0.44 |
| Goodness of fit (R<sup>2</sup>) | 0.35 | 0.27 | 0.43 |

<sup>a</sup> Within-village Z score
<sup>b</sup> Mean of population-level Z scores within a village
<sup>c</sup> Population-level Z score, coefficient calculated from posterior samples of a+b

**Table S7:** Model summary – BMI.

|  | Mean | Lower<br>95% CI | Upper<br>95% CI |
| --- | --- | --- | --- |
| Intercept | 0.10 | 0.02 | 0.18 |
| Gini | -0.01 | -0.08 | 0.06 |
| Sex=male | 0.03 | -0.03 | 0.10 |
| Relative household wealth<sup>a</sup> | -0.01 | -0.04 | 0.02 |
| Age | 0.10 | 0.07 | 0.14 |
| Mean community-level wealth<sup>b</sup> | 0.09 | -0.10 | 0.29 |
| Distance to San Borja | -0.09 | -0.16 | -0.01 |
| Community Size | -0.01 | -0.09 | 0.06 |
| Absolute household wealth<sup>c</sup> | 0.09 | -0.12 | 0.28 |
| Goodness of fit (R<sup>2</sup>) | 0.72 | 0.72 | 0.73 |

<sup>a</sup> Within-village Z score
<sup>b</sup> Mean of population-level Z scores within a village
<sup>c</sup> Population-level Z score, coefficient calculated from posterior samples of a+b

**Table S8:**
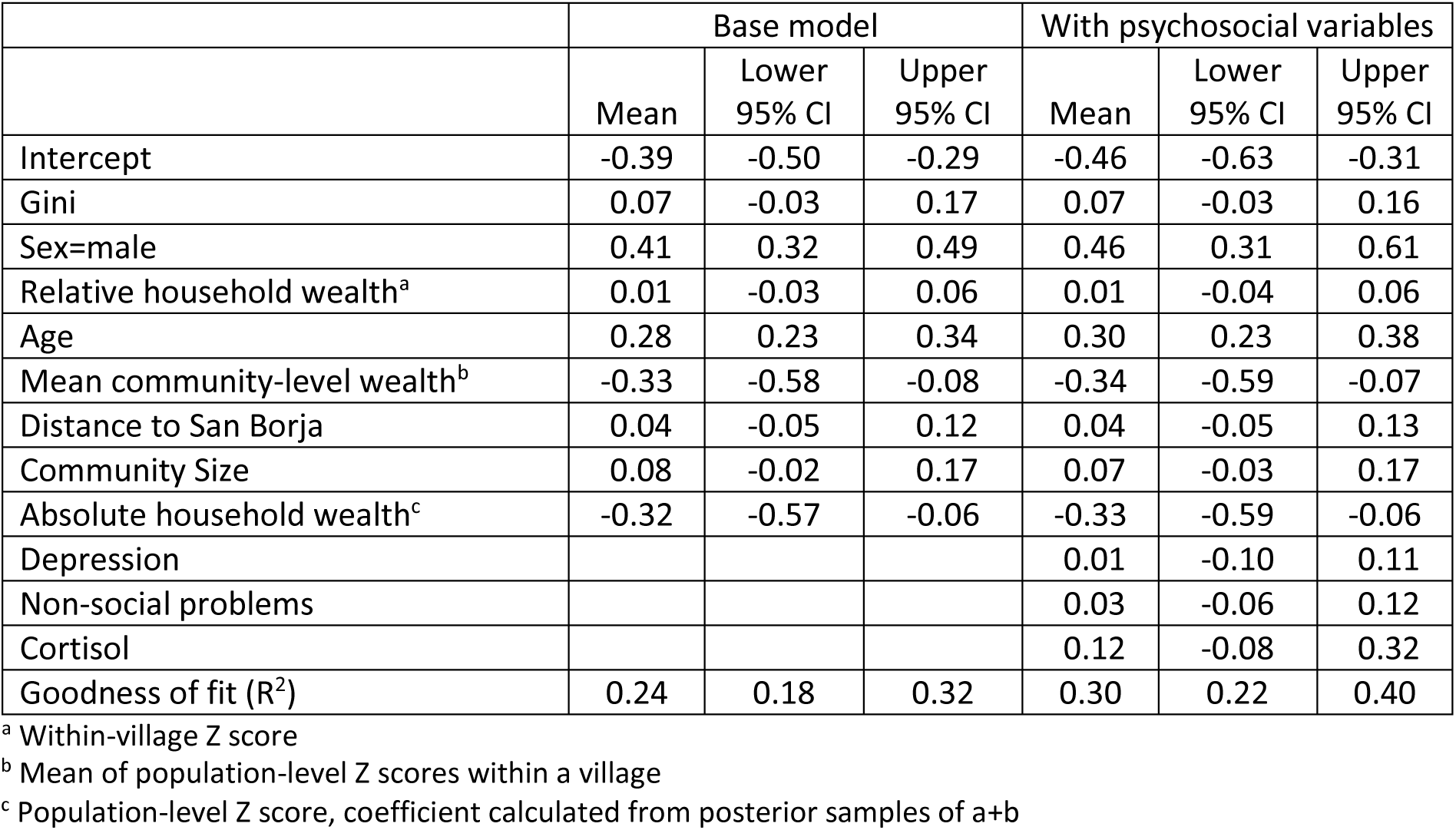
Model summary – Systolic blood pressure.

|  | Base model |  |  | With psychosocial variables |  |  |
| --- | --- | --- | --- | --- | --- | --- |
|  | Mean | Lower<br>95% CI | Upper<br>95% CI | Mean | Lower<br>95% CI | Upper<br>95% CI |
| Intercept | -0.39 | -0.50 | -0.29 | -0.46 | -0.63 | -0.31 |
| Gini | 0.07 | -0.03 | 0.17 | 0.07 | -0.03 | 0.16 |
| Sex=male | 0.41 | 0.32 | 0.49 | 0.46 | 0.31 | 0.61 |
| Relative household wealth<sup>a</sup> | 0.01 | -0.03 | 0.06 | 0.01 | -0.04 | 0.06 |
| Age | 0.28 | 0.23 | 0.34 | 0.30 | 0.23 | 0.38 |
| Mean community-level wealth<sup>b</sup> | -0.33 | -0.58 | -0.08 | -0.34 | -0.59 | -0.07 |
| Distance to San Borja | 0.04 | -0.05 | 0.12 | 0.04 | -0.05 | 0.13 |
| Community Size | 0.08 | -0.02 | 0.17 | 0.07 | -0.03 | 0.17 |
| Absolute household wealth<sup>c</sup> | -0.32 | -0.57 | -0.06 | -0.33 | -0.59 | -0.06 |
| Depression |  |  |  | 0.01 | -0.10 | 0.11 |
| Non-social problems |  |  |  | 0.03 | -0.06 | 0.12 |
| Cortisol |  |  |  | 0.12 | -0.08 | 0.32 |
| Goodness of fit (R<sup>2</sup>) | 0.24 | 0.18 | 0.32 | 0.30 | 0.22 | 0.40 |

<sup>a</sup> Within-village Z score
<sup>b</sup> Mean of population-level Z scores within a village <sup>c</sup> Population-level Z score, coefficient calculated from posterior samples of a+b

**Table S9:** Model summary – Diastolic blood pressure.

|  | Base model |  |  | With psychosocial variables |  |  |
| --- | --- | --- | --- | --- | --- | --- |
|  | Mean | Lower 95% CI | Upper 95% CI | Mean | Lower 95% CI | Upper 95% CI |
| Intercept | -0.28 | -0.43 | -0.13 | -0.35 | -0.55 | -0.15 |
| Gini | 0.08 | -0.05 | 0.23 | 0.08 | -0.05 | 0.22 |
| Sex=male | 0.25 | 0.16 | 0.34 | 0.31 | 0.16 | 0.49 |
| Relative household wealth<sup>a</sup> | -0.02 | -0.07 | 0.03 | -0.02 | -0.07 | 0.03 |
| Age | 0.23 | 0.18 | 0.29 | 0.25 | 0.18 | 0.32 |
| Mean community-level wealth<sup>b</sup> | -0.36 | -0.71 | -0.01 | -0.34 | -0.70 | 0.01 |
| Distance to San Borja | 0.00 | -0.13 | 0.12 | 0.01 | -0.12 | 0.13 |
| Community Size | 0.07 | -0.07 | 0.22 | 0.07 | -0.09 | 0.23 |
| Absolute household wealth<sup>c</sup> | -0.38 | -0.74 | -0.04 | -0.36 | -0.72 | 0.00 |
| Depression |  |  |  | -0.01 | -0.11 | 0.09 |
| Non-social problems |  |  |  | 0.04 | -0.05 | 0.13 |
| Cortisol |  |  |  | 0.12 | -0.11 | 0.33 |
| Goodness of fit (R<sup>2</sup>) | 0.22 | 0.16 | 0.29 | 0.27 | 0.19 | 0.37 |

<sup>a</sup> Within-village Z score <sup>b</sup> Mean of population-level Z scores within a village <sup>c</sup> Population-level Z score, coefficient calculated from posterior samples of a+b

**Table S10:**
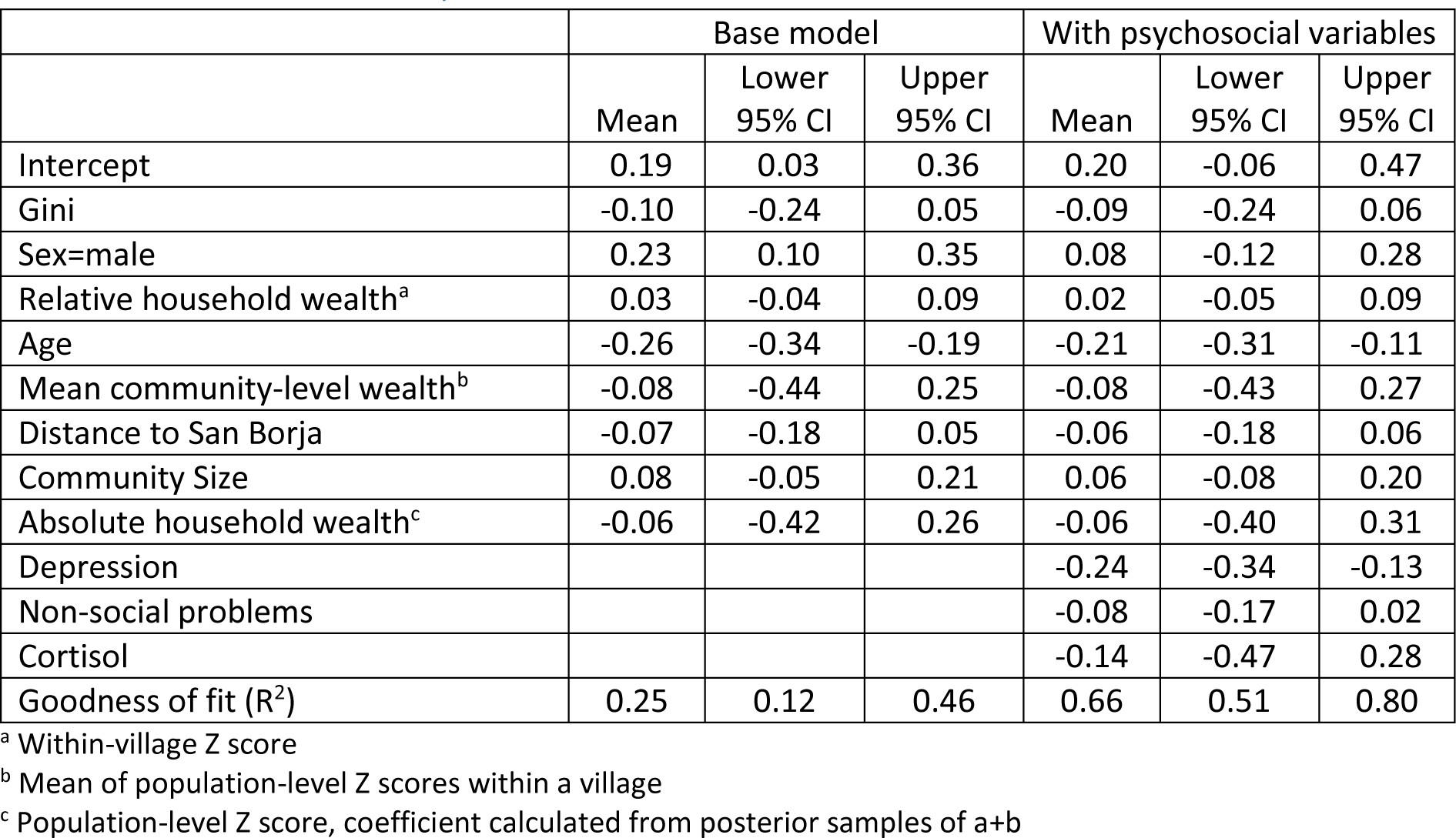
Model summary – Self-rated health.

**Table S11:** Model summary – Total morbidity.

|  | Base model |  |  | With psychosocial variables |
| --- | --- | --- | --- | --- |

|  | Mean | Lower<br>95% CI | Upper<br>95% CI | Mean | Lower<br>95% CI | Upper<br>95% CI |
| --- | --- | --- | --- | --- | --- | --- |
| Intercept | 0.11 | -0.10 | 0.34 | 0.21 | -0.02 | 0.44 |
| Gini | -0.07 | -0.26 | 0.10 | -0.07 | -0.25 | 0.11 |
| Sex=male | -0.14 | -0.20 | -0.08 | -0.12 | -0.25 | 0.00 |
| Relative household wealth<sup>a</sup> | -0.03 | -0.07 | 0.01 | -0.03 | -0.07 | 0.01 |
| Age | 0.34 | 0.31 | 0.37 | 0.32 | 0.28 | 0.36 |
| Mean community-level wealth<sup>b</sup> | -0.04 | -0.51 | 0.43 | -0.07 | -0.54 | 0.39 |
| Distance to San Borja | 0.10 | -0.08 | 0.27 | 0.11 | -0.07 | 0.28 |
| Community Size | -0.06 | -0.27 | 0.15 | -0.04 | -0.25 | 0.18 |
| Absolute household wealth<sup>c</sup> | -0.07 | -0.54 | 0.39 | -0.10 | -0.58 | 0.35 |
| Depression |  |  |  | 0.12 | 0.04 | 0.19 |
| Non-social problems |  |  |  | -0.10 | -0.16 | -0.04 |
| Cortisol |  |  |  | -0.06 | -0.25 | 0.12 |
| Goodness of fit (R<sup>2</sup>) | 0.38 | 0.36 | 0.41 | 0.42 | 0.39 | 0.47 |

<sup>a</sup> Within-village Z score
<sup>b</sup> Mean of population-level Z scores within a village
<sup>c</sup> Population-level Z score, coefficient calculated from posterior samples of a+b

**Table S12:** Model summary – Infections. Note that unlike in previous models, these estimates are on the logit scale

|  | Base model |  |  | With psychosocial variables |  |  |
| --- | --- | --- | --- | --- | --- | --- |
|  | Mean | Lower 95% CI | Upper 95% CI | Mean | Lower 95% CI | Upper 95% CI |
| Intercept | -1.74 | -2.40 | -1.03 | -1.91 | -2.73 | -1.14 |
| Gini | -0.47 | -1.04 | 0.06 | -0.51 | -1.07 | 0.04 |
| Sex=male | -0.48 | -0.69 | -0.25 | -0.50 | -0.95 | -0.07 |
| Relative household wealth<sup>a</sup> | -0.02 | -0.17 | 0.12 | -0.02 | -0.17 | 0.13 |
| Age | 0.57 | 0.45 | 0.69 | 0.63 | 0.47 | 0.82 |
| Mean community-level wealth<sup>b</sup> | 0.33 | -0.82 | 1.48 | 0.29 | -0.93 | 1.51 |
| Distance to San Borja | -0.20 | -0.68 | 0.29 | -0.20 | -0.72 | 0.31 |
| Community Size | -0.34 | -0.95 | 0.30 | -0.38 | -1.04 | 0.28 |
| Absolute household wealth<sup>c</sup> | 0.31 | -0.82 | 1.48 | 0.27 | -0.97 | 1.47 |
| Depression |  |  |  | -0.20 | -0.52 | 0.10 |
| Non-social problems |  |  |  | -0.01 | -0.25 | 0.24 |
| Cortisol |  |  |  | 0.18 | -0.51 | 0.87 |
| Goodness of fit (R<sup>2</sup>) | 0.31 | 0.27 | 0.34 | 0.33 | 0.28 | 0.38 |

<sup>a</sup> Within-village Z score
<sup>b</sup> Mean of population-level Z scores within a village
<sup>c</sup> Population-level Z score, coefficient calculated from posterior samples of a+b

**Table S13:** Model summary – Respiratory illness. Note that unlike in previous models, these estimates are on the logit scale

|  | Base model |  |  | With psychosocial variables |  |  |
| --- | --- | --- | --- | --- | --- | --- |
|  | Mean | Lower 95% CI | Upper 95% CI | Mean | Lower 95% CI | Upper 95% CI |
| Intercept | -0.92 | -1.34 | -0.50 | -1.31 | -2.18 | -0.63 |
| Gini | 0.31 | -0.06 | 0.70 | 0.35 | -0.07 | 0.82 |
| Sex=male | -0.06 | -0.25 | 0.11 | 0.48 | 0.01 | 1.09 |
| Relative household wealth<sup>a</sup> | -0.13 | -0.24 | -0.01 | -0.14 | -0.28 | -0.01 |
| Age | -0.54 | -0.65 | -0.44 | -0.54 | -0.78 | -0.31 |
| Mean community-level wealth<sup>b</sup> | 0.07 | -0.82 | 1.00 | 0.06 | -0.95 | 1.07 |
| Distance to San Borja | 0.22 | -0.15 | 0.58 | 0.25 | -0.15 | 0.66 |
| Community Size | 0.15 | -0.29 | 0.56 | 0.18 | -0.30 | 0.68 |
| Absolute household wealth<sup>c</sup> | -0.06 | -0.95 | 0.85 | -0.08 | -1.11 | 0.92 |
| Depression |  |  |  | 0.19 | -0.16 | 0.55 |
| Non-social problems |  |  |  | -0.16 | -0.46 | 0.14 |
| Cortisol |  |  |  | 0.96 | 0.26 | 1.78 |
| Goodness of fit (R<sup>2</sup>) | 0.23 | 0.20 | 0.27 | 0.33 | 0.24 | 0.45 |

<sup>a</sup> Within-village Z score
<sup>b</sup> Mean of population-level Z scores within a village
<sup>c</sup> Population-level Z score, coefficient calculated from posterior samples of a+b

**Table S14:**
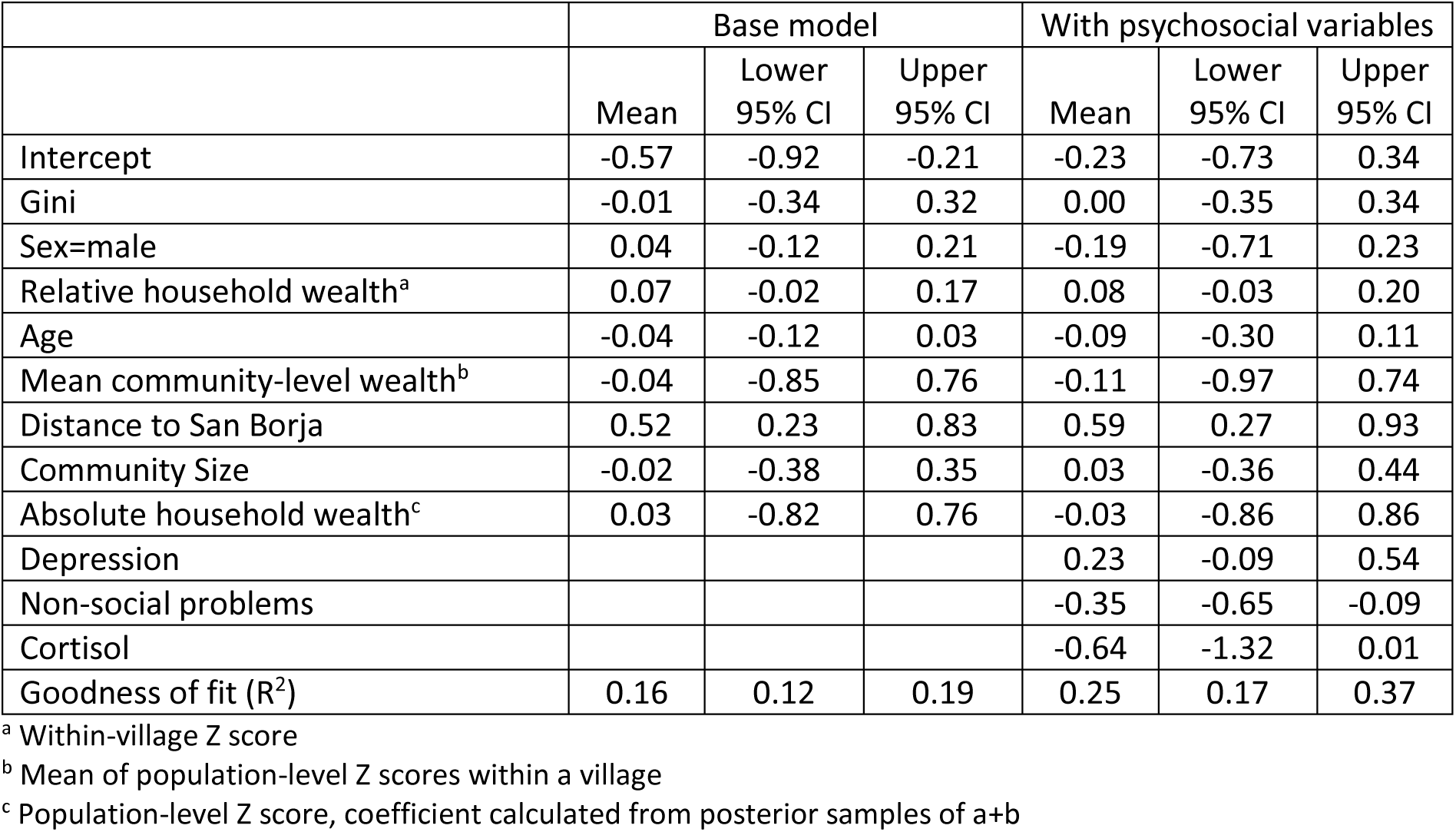
Model summary – Gastrointestinal illness. Note that unlike in previous models, these estimates are on the logit scale

**Table S15:** Overview of exploratory interaction effects. Interactions were added in a stepwise forward selection procedure. That is, if the addition of any interaction (Sex * Wealth, Sex * Gini, Gini * Wealth) improved the expected log pointwise predictive density (ELPD) compared to the base model, this interaction was included. The respective model was then compared to models with two interactions, etc. Interpretations are based on posterior probability supporting an interaction and graphical inspection

| Outcome | Interactions added | Interpretation |
| --- | --- | --- |
| Depression | Sex * Wealth | Wealth-depression association only found in men |
| Social conflicts | - |  |
| Labor partners | Gini * Wealth | In unequal places, wealthy people have more labor partners; in equal places, poor people have more |
| Non-social problems | - |  |
| Cortisol | Sex * Gini | Gini-cortisol association only seen in men |
| Body Mass Index | -<sup>a</sup> |  |
| Systolic blood pressure | - |  |
| Diastolic blood pressure | Gini * Wealth | Gini-BP association much stronger in richer people |
|  | Sex * Gini | Gini-BP association only seen in women |
| Self-rated health | - |  |
| Total morbidity | - |  |
| Infections | Sex * Gini | Women slightly higher than men in more equal places |
| Respiratory illness | Sex * Wealth | Wealth-health association only seen in women |
| Gastrointestinal illness | Sex * Gini | Gini effect slightly stronger for men, weak support |

<sup>a</sup> Note that stepwise model selection was not computationally feasible for BMI because robust comparisons took days to weeks to run. A comparison of just the base model and a model with *all* possible interactions favored the base model

